# PROBLEM BASED LEARNING APPLIED TO PRACTICE IN MEDICAL SCHOOLS IN BRAZIL: A MINI-SYSTEMATIC REVIEW

**DOI:** 10.1101/19007955

**Authors:** Frederico Alberto Bussolaro, Claudine Thereza-Bussolaro

## Abstract

**Background:** Active learning is a well-established educational methodology in medical schools worldwide, although its implementation in Brazilian clinical settings is quite challenging. The objective of this study is to review the literature in a systematic manner to find and conduct a reflective analysis of how problem-based learning (PBL) has been applied to clinical teaching in medical schools in Brazil.

**Material & methods:** A systematic literature search was conducted in three databases. A total of 250 papers related to PBL in Brazilian medical schools were identified through the database searches. Four studies were finally selected for the review.

**Results:** Four fields of medicine were explored on the four selected papers: gynecology/family medicine, medical semiology, psychiatry, and pediatrics. Overall, all the papers reported some level of strategic adaptability of the original PBL methodology to be applied in the Brazilian medical school’s curricula and to the peculiar characteristics specific to Brazil.

**Conclusion:** PBL application in Brazilian medical schools require some level of alteration from the original format, to better adapt to the characteristics of Brazilian students’ maturity, health system priorities and the medical labor market.

## INTRODUCTION

Problem-based learning, a constructivist method, was introduced to the medical curriculum in 1969 (1) to help students become effective solvers of biomedical problems and to foster attitudes leading to behavior as responsible physicians and scientists in relation to their patients.

The constructivism view of learning has evolved since the beginning of its concept in 1960. The need for more critical humanist and reflective ethical health-related professionals, promoted the tendency to use “active” methods. The concept of an active methodology is that the students must be active during their learning process, meaning that books, slides, online presentations, and lectures are considered passive and non-constructivist venues of teaching; whereas, interactive games, hands-on activities, and group discussions are classified as constructivist i.e. active venues. A systematic review(2) analyzed the effects of PBL on the future physician and conclude that PBL during medical school has a positive effect on physician competency.

Active learning emphasizes that knowledge cannot be transmitted but needs to be constructed.(3) The active methodology has been spread worldwide by the discovery-based learning methodology (DBL). Both students and professors are keen on the DBL methodology.(2) DBL is a constructivist based approach in education i.e. a “learn by doing” method in which the students are provided with the materials to find the answers.

The challenges in medicine are related to patient safety concerns, operating room efficiencies, and duty-hour restrictions.(3) The traditional apprenticeship-style “see one, do one, teach one” has significant disadvantages, in today’s pressured academic environment where research productivity and clinical and operating room efficiencies are prioritized over teaching time.(4)

Similarly challenging, PBL has a rigorous and highly structured practical teaching-learning process in which the student learns while trying to understand or solve a problem.(5) PBL is structured in “problem, solution, practice, research, questioning, realism, originality and integration”(6), therefore its implementation in clinical settings requires more time, higher costs and more training of teachers, and is affected not only by the educational aspects but for the unique priorities of health care environments as well.(7, 8)

Brazilian program curricula abide by the 2001 national guidelines for medical courses of the ministry of education. The guideline requires that 35% of the total workload of the medical curricula is dedicated to clerkship during the last two years of the program. Additionally, the guidelines require that students be exposed to patients of different levels of clinical complexity, difficulty, and assistance. Therefore, the guidelines strongly encourage the use of PBL and suggests that students see patients early on in their medical program.(9)

PBL methodology has been introduced to medical schools in Brazil since 1997.(10) This study aims to review the literature in a systematic manner to find and conduct a reflective analysis about how PBL has been applied to clinical teaching in Brazilian medical schools.

## MATERIAL AND METHODS

This is a study based on the mini-systematic literature review on PBL methodology adopted in Brazil’s medical schools.

### Protocol and registration

This review followed as much as possible the preferred reporting items for systematic review and meta-analyses (PRISMA). We did not register the protocol of this review.

### Eligibility Criteria

The identification of the studies was based on the research question: what field of medicine has been applying PBL methodology in medical schools in Brazil?

#### Inclusion criteria

This review included retrospective and prospective studies concerning active learning, specifically the PBL methodology, adopted in medical schools in Brazil.

#### Exclusion criteria

Review articles, and articles in which the methodology was applied to fields other than medicine. Articles that did not present information related to the clinical applicability of the PBL methodology were excluded.

### Search strategy

Electronic databases searched were LILACS, MEDLINE and PsycINFO

The search approach was developed through consultation with medical professors and reviewing articles related to the topic. Appropriate keyword, truncation and word combinations were adapted.

### Data search

Three databases were searched LILACS, PsycINFO and MEDLINE, plus a manual search were conducted from the reference lists of the included articles. The literature search was conducted on July 2,2019 with the words “PBL” OR “Problem based learning” AND “Brazil*” AND “medical school” OR “medicine”. For the LILACS search words in Portuguese were added. A total of 250 papers were found as follow: LILACS: 113 papers; PsycINFO: 84 papers; MEDLINE: 53 papers.

After duplicates were removed, a total of 238 articles were screened by title and abstract. Finally a total of four articles met our eligibility criteria. A flowchart of the selection can be seen in figure 1.

**Figure 1.**
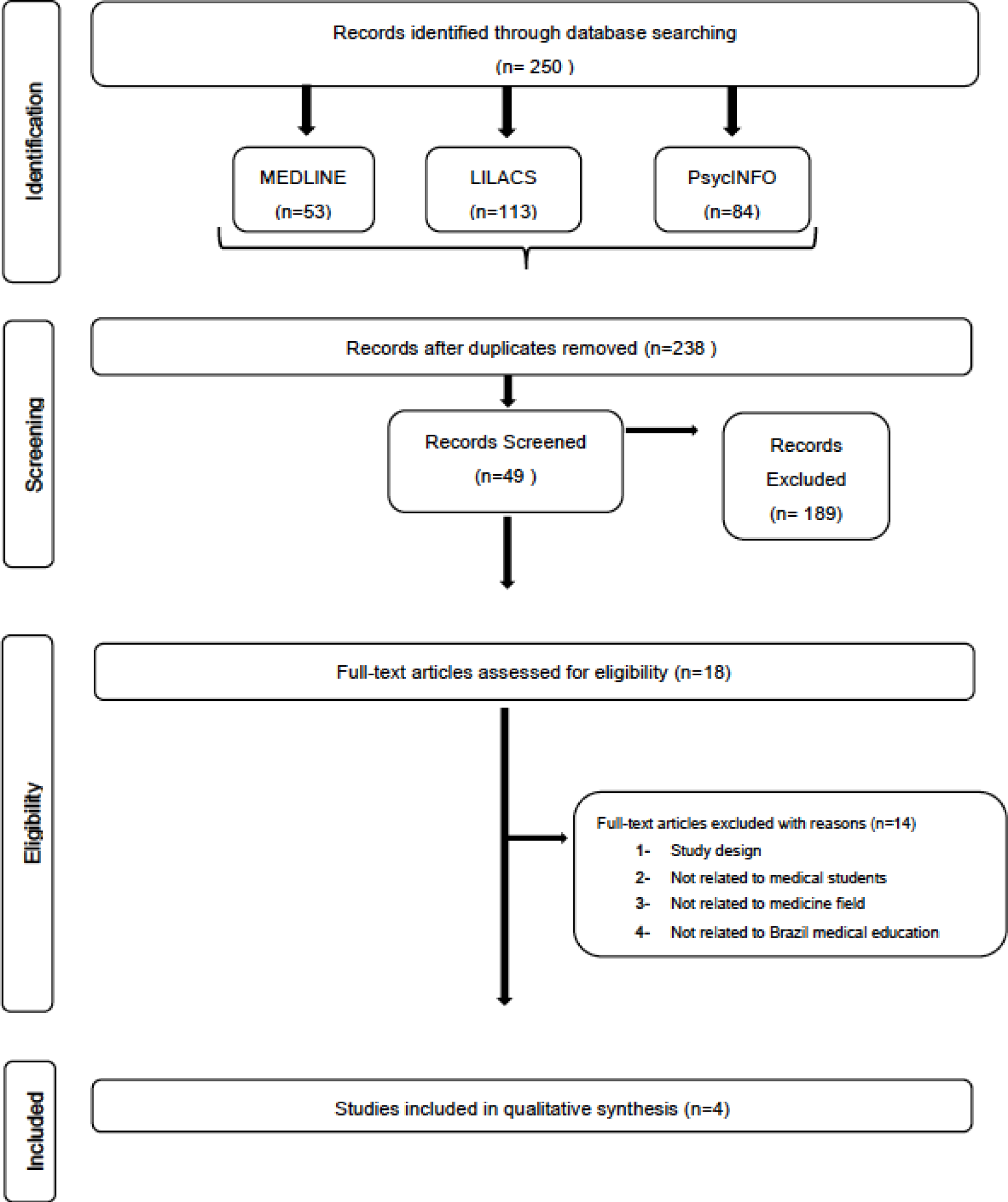
Flowchart of the review

### Study selection and Eligibility

All abstracts identified during the database search were screened by two independent reviewers (F.A.B. and C.T.B). Potentially relevant abstracts were then selected for full article evaluation by the same two independent reviewers. Any selection discrepancies were resolved through discussion between the two reviewers.

### Strategy for data extraction and selection

The main outcome was applicability of PBL as the teaching methodology within the clinical setting in medicine. The organization of the results was based on the medical clinical field.

## RESULTS

A total of 250 papers were found and, after applying the inclusion and exclusion criteria, 4 papers (10-13) were definitely selected for this review. Table 1 shows the summary of findings.

**Table 1.** Literature Review summary of Findings.

| Author | Year | State(Brazil) | Field of application | Students (n) | Year | Integrated practic | Objective |
| --- | --- | --- | --- | --- | --- | --- | --- |
| Paulin & Poças | 2009 | São Paulo | Psychiatric | 80 | 5th | yes | experience report |
| Prado et al. | 2011 | Pernambuco | Paediatric | 72 | 5th | yes | compare educational effectiveness of ward rounds conducted whit two different methodologies |
| Bestetti et al. | 2014 | São Paulo | Gynecology,internal medicine, family physician | N/R | 5th year (8-10 semester) | yes | experience report |
| Dias-Lima et al. | 2019 | Bahia | Medical Semiology | N/R | 2 & 3rd semester | yes | experience report |

n, number; NR, Not reported

### Fields of application

Four fields of medicine were explored on the selected papers: gynecology/family medicine, medical semiology, psychiatry and pediatrics.

#### Gynecology and family medicine

the authors believe PBL is a worthwhile approach to teach Brazilian medical students. Bestetti et al. (11) reported a coordinate mix (tutoring, medical skills and primary care) in which students are trained in clinics and labs after the tutorial part of PBL; and, at the end of the process, students are expected to solve patient health problems.

#### Aggression and Defense Mechanism(Medical Semiology)

the authors Dias-Lima et al. (12) observed a positive impact on student training and interest in the subjects taught.

#### Psychiatry

Paulin & Poças (10) report on the implementation of the PBL methodology to a psychiatry internship. The authors noticed improvement on the students’ knowledge regarding psychiatry. Another observation was proper patient referral from former students who had been taught under the same methodology.

#### Paediatric Clinical teaching-ward rounds

Prado et al. assessed the contribution of two different methodologies - traditional methodology and active methodology - in improving students’ knowledge. The study was performed on three phases, and true/false pre-test assessment and post-test assessments were applied. Prado et al. (13) study found that students’ knowledge on pneumonia and diarrhea improved 3.5 more when the active methodology was applied.

## DISCUSSION

During this mini-systematic review, four studies reported the use of PBL methodology in Brazilian medical schools (10-13), in hospitals, internships, and simulation labs.

It should be explained that PBL was introduced in Canada in a three-year undergrad curriculum rather than the traditional four-year program.(1) The traditional four-year program has four-month of break in the summer, while the three-year PBL curriculum has only one-month therefore students have 112 weeks of academic activity plus 12 weeks of elective time as seen in figure 2. PBL has four phases:

- Phase I lasts 14 weeks and is dedicated to normal structure and function;
- Phase II lasts six weeks and is dedicated to abnormal biological mechanisms;
- Phase III lasts 40 weeks and is dedicated to abnormal structure and function;
- Phase IV is dedicated to the clerkships in four interchangeable blocks of: family medicine practice, surgery and psychiatry, pediatrics, and obstetrics and gynecology.

**Figure 2.**
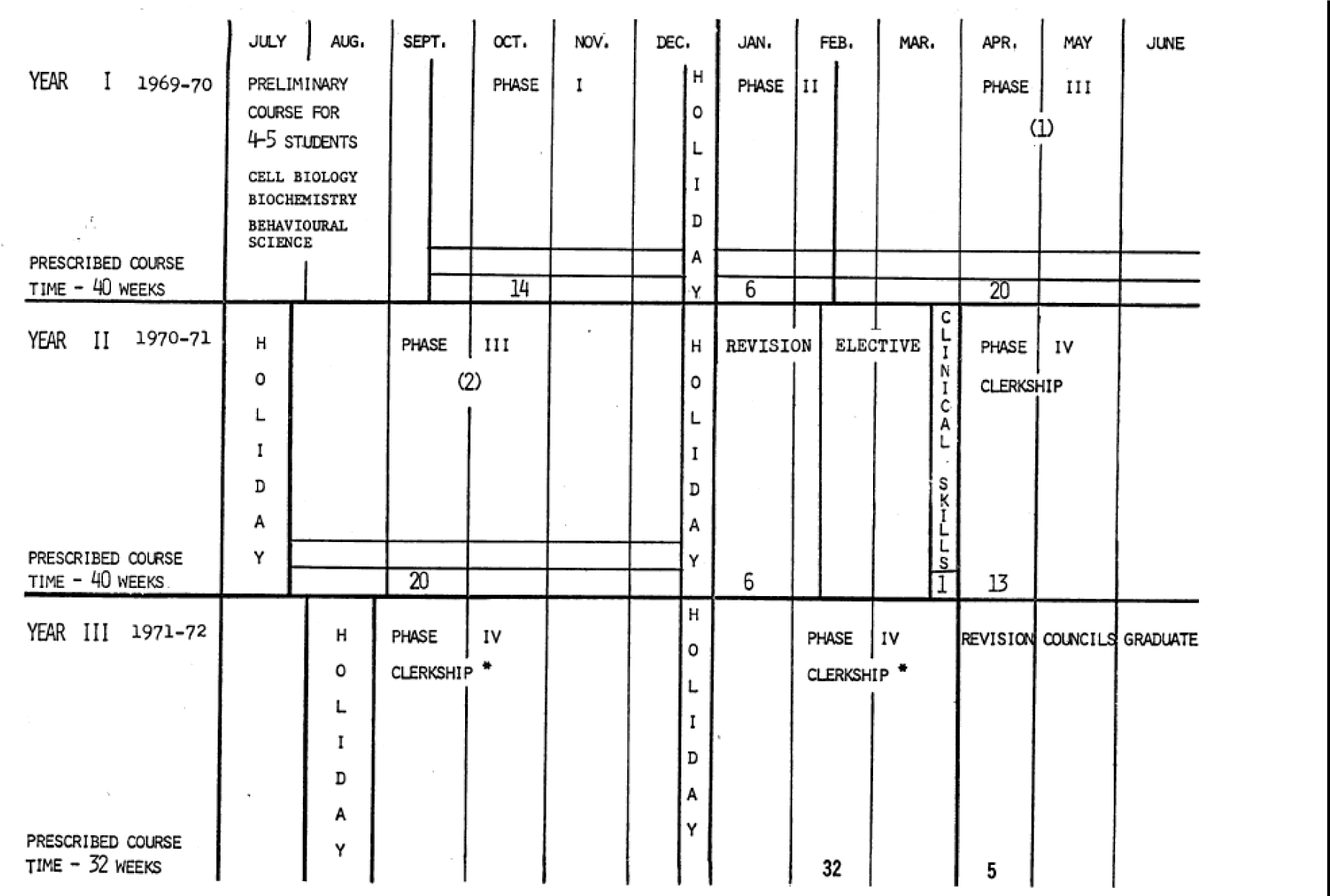
Outline of the three-year undergrad medical curriculum proposed by McMaster University, including six weeks of elective time (retrieved from Spaulding, 1969, page 660) (1)

Throughout Phase I to Phase III students also have a horizontal program (one hour per day) dedicated to problems related to professional attitudes and ethics, abnormal behavior, biomedical statistics, epidemiology, and rehabilitation medicine. Furthermore, students have two six-week periods of elective experiences arranged by faculty members, and before entering the clerkship students have a week of clinical skills in history taking, examination, and planning of further investigation and treatment.

Additionally, it should be stressed that the requirements for a student to be admitted to medical schools are different between Canada and Brazil. In Canada, the PBL program was proposed for students who already had at least three years in behavioral science, biochemistry, and cell biology, and students should preferably have completed a degree program. In Brazil, the requirement to enter a medical school is to pass in an exam which evaluates student’s knowledge in languages, codes and related technologies, as well as various subjects such as Portuguese, History, Geography, Mathematics, Physics, Chemistry and Biology and a written essay. Therefore the maturity and the academic backgrounds are different between students in the first year of medical school, factors that can compromise the success of PBL in medical schools in Brazil.

The maturity issue among Brazilian students has been observed. Accordingly (11), Brazilian medical students lack maturity and confidence to excel in a PBL approach, leading them to be anxious and insecure about their short-term learning in the context of PBL methodology.(11)

Nonetheless, the Brazilian medical school curriculum has been linked wherever possible with the PBL methodology. Bestetti et al. (11) relate the importance of cultural and structural context in implementing PBL in Brazilian medical schools. In Brazil, the implementation of the PBL methodology is said to be affected by the medical labor market and the health system as well as the educational aspect.(11)

Factors such as time constraints, clinical flows, and diversity of clinical problems impact teaching conditions.(8) Barrows, 1986 stated the PBL method requires more time, costs more, and requires more effort from teachers compared to the lecture-based methods. Accordingly, an effective PBL method requires a quality of tutorial skills, meaning that the teacher or tutor has to guide the students to consider all the steps in the reasoning of the problem. If this fails, the objectives of the PBL are compromised.(14) Another issue that can compromise the use of PBL in the clinical teaching setting is that clinical teachers are always engaged in a dual process: clinical reasoning to care for patients, and educational reasoning to teach patients.(8)

Couto et al.(15), analyzed Brazilian medical students perceptions of subject-matter expertise among PBL facilitators, and they concluded that PBL facilitator experts in the topic are essential to the learning process. Similarly, in Canada the birthplace of PBL, studies (16- 18) have shown that for the success of the PBL methodology, professors should be able to facilitate groups and be knowledgeable about the content, and at the same time students need to be willing to participate in peer teaching and be supportive of the group leaning process. It is expected that the professors have expertise in their field.(16-18)

The McMaster University medical school that initially proposed the PBL curriculum program has also undergone modifications since its first implementation over 30 years ago. They maintained the philosophy but changed the curriculum in 2005, implementing the Compass (Concept-oriented, multidisciplinary, problem-based, practice for transfer, simulations in clerkship, streaming). The major changes were in student evaluations, the roles of the tutor, and the number and purpose of educational sessions.(17) Figure 3 shows the outline of the three-year Compass undergrad curriculum currently in place at McMaster University.

**Figure 3.**
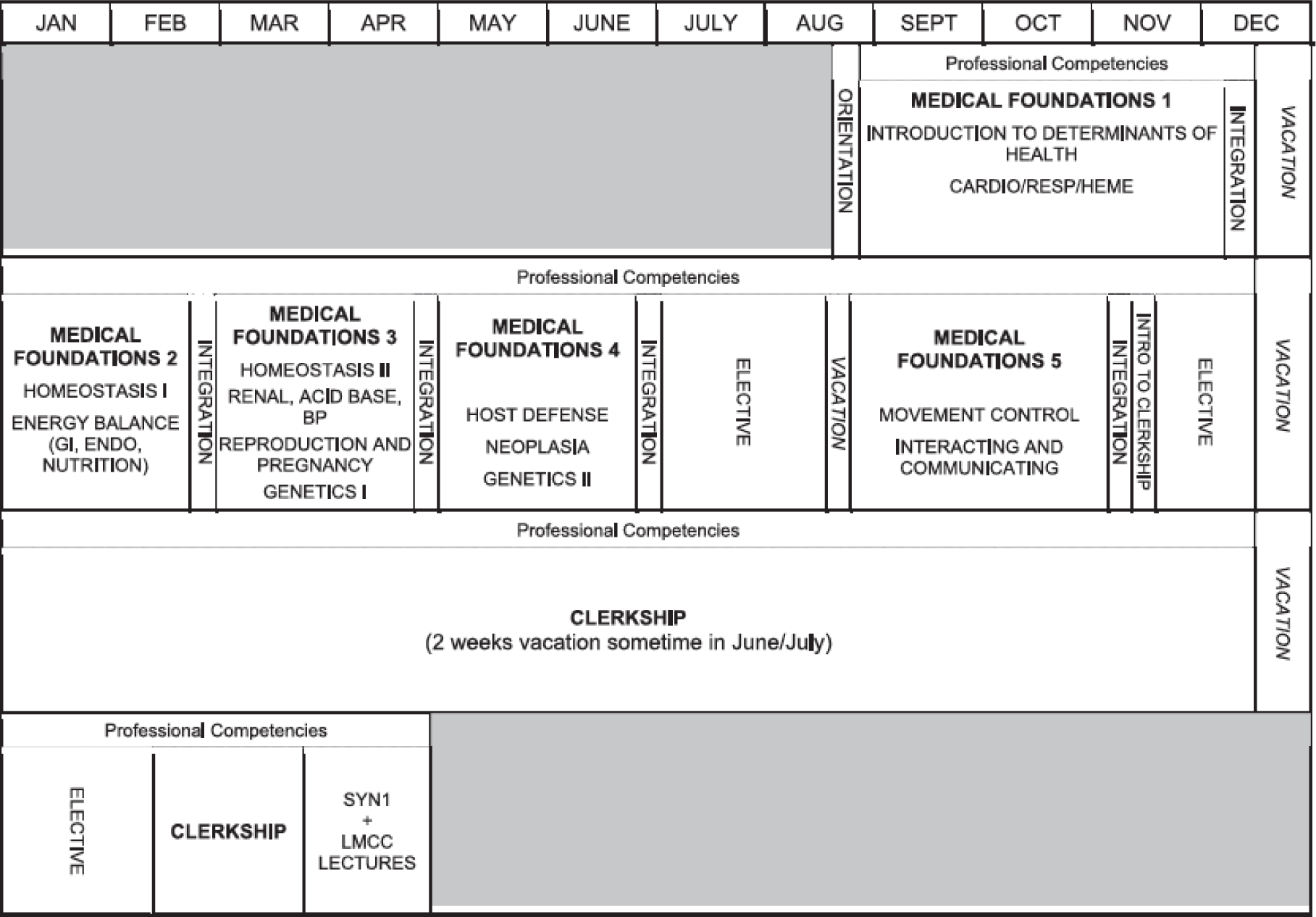
Outline of the three-year Compass undergraduate MD curriculum retrieved from Neville & Norman, page 373(17)

In our review, we found that Bestetti (11) also related the importance of subject-matter expertise tutors to minimize time preparation and maximize student confidence. Additionally, they invest in courses and preparation of tutors within the practice and theoretical content with the PBL methodology. In contrast, most teachers in Brazil are more familiar with the traditional lecture-based methodology rather than the student-centered approach used in the PBL methodology.

Dias-Lima et al.(12) reported a positive impact of the student training part of their program. Although, their program has adapted the PBL method with educational tools to integrate the apprenticeship of disciplines like microbiology, parasitology, immunology, and pathology in a clinical context. The drawback of the study is the absence of inclusion of patients in the discipline taught; they used videos to simulate a clinical environment.

Similarly, Lucchetti et al.(19), describe the experience of active methodology, including simulation, to teach geriatrics and observed improvement in medical student’s knowledge, attitudes, and skills. Geriatrics is a very challenge area to teach in comparison to other areas where the presence of technology and procedures turns them into more attractive areas for students. Although the paper addressed the use of active methodology, it did not specify the use of PBL; therefore, it was excluded from our analyses.

In the pediatric field (13), researchers selected six patients from the emergency room with three patients diagnosed with diarrhea and three with pneumonia. Seventy-two students were submitted to a test prior to assessing the patients, after that students participated in the academic ward round conducted by a staff member previously trained in both methodologies, TM (traditional method) and AM (active methodology). Lastly, students were submitted to a second cognitive test 48 hours after the ward round. The authors concluded that AM showed better results on students’ knowledge acquisition perception.

In Brazil, learning in the internship phase is done with hospitalized patients in hospital beds, basic units, specialized outpatient clinics, or emergency rooms. These situations usually require immediate care to the patient in the hospital, which limit studies and tutorial discussions. To eliminate or even clear some of the limitations, the studies by Paulin & Pocas (10) and Prado et al.(13), demonstrated adaptations/models of hospital and outpatient care focused on learning with PBL, situations that need adaptations/modifications in the healthcare system that assists patients, allows the creation of moments for studies and discussion of cases without impairing the quality of the care of the patients. Both teachers and students need to adapt with a greater sense of responsibility in preparing with studying prior to care and thus aiming to reduce insecurities. Students can increase their dedication to practices in laboratories/semiology activities; teachers can widen the search to bring tools that allow students to be motivated. Teachers need to prepare and understand the new profile of these students, who ask more questions, due to the broad access to information available on the internet and from varied sources which are not always scientific.

PBL in medical education has been presented, for the most part, as a prospect for improving the skills of physicians trained by this active methodology. However, these studies have not shown a convincing improvement in the knowledge and performance gains of trained medical professionals using this methodology as compared to the traditional methods used in most Brazilian medical schools until the end of the last century.

## CONCLUSION

PBL application in medical schools in Brazil undergo some level of alteration from the original format, to better adapt to the peculiarities of Brazilian student maturity, health system priorities, and the medical labor market.

In Brazil, more studies on PBL methodology need to be conducted to evaluate the student knowledge acquisition through active learning methodology.

## Data Availability

data sharing is not applicable to this article as no new data were created or analyzed in this study.

